# TROPONIN IS INDEPENDENTLY ASSOCIATED WITH DEATH IN PATIENTS WITH COVID: A RETROSPECTIVE STUDY

**DOI:** 10.1101/2021.12.23.21268005

**Authors:** Vijay Shyam-Sundar, Dan Stein, Martina Spazzapan, Andrew Sullivan, Cathy Qin, Victor Voon

## Abstract

**Objective:** We performed a single-centre retrospective observational study investigating the association between troponin positivity in patients hospitalised with COVID-19 and increased mortality in the short term.

**Methods:** All adults admitted with swab-proven RT-PCR COVID-19 to Homerton University Hospital (HUH) from 04.02.20 to 30.04.20 were eligible for inclusion.

We retrospectively analysed demographic and biochemical data collected from the physical and electronic patient records according to the primary outcome of death at 28 days during hospital admission.

Troponin positivity was defined above the upper limit of normal according to our local laboratory assay (>15.5ng/l for females, >34 ng/l for males). Univariate and multivariate logistical regression analyses were performed to evaluate the link between troponin positivity and death.

**Results:** Mean length of stay for all 402 hospitalised COVID-19 patients at HUH was 9.1 days (SD 12.0). Mean age was 65.3 years for men compared to 63.8 years for women. A chi-squared test showed that survival of COVID-19 patients was significantly higher in those with a negative troponin (p = 3.23 ×10^−10^) compared to those with a positive troponin. In the multivariate logistical regression, lung disease, age, troponin positivity and CPAP were all significantly associated with death, with an AUC of 0.8872, sensitivity of 0.9004 and specificity of 0.6292 for the model. Within this model, troponin positivity was independently associated with short term mortality (OR 3.23, 95% CI 1.53-7.16, p=0.00278).

**Conclusions:** We demonstrated an independent association between troponin positivity and increased short-term mortality in COVID-19 in a London district general hospital.

**Key Questions:** *What is already known about this subject?:* An elevated Troponin is associated with increased mortality. Troponin is known to be elevated in some patients who test positive for COVID-19 infection.

*What does this study add?:* This study shows an association between an elevated troponin in hospitalised COVID-19 patients and increased short-term mortality.

*How might this impact on clinical practice?:* Troponin is a readily available, easy to measure biomarker which can be used to predict the severity of COVID-19 illness and could aid prognostication in hospital.

## Introduction

The COVID-19 pandemic has so far resulted in over 200 million recorded infections and over 4 million deaths worldwide (1). A significant proportion of patients develop acute respiratory distress syndrome (ARDS), require hospitalisation and ventilatory support with subsequent morbidity and mortality. Early data from Wuhan, first outlined the association of COVID-19 and myocardial injury exhibited by elevated cardiac biomarkers (2–4). This has subsequently been replicated in various studies and meta-analyses from around the world (5).

The mechanisms for myocardial injury in COVID-19 are not fully understood. A combination of systemic hypoxia, cytokine storm, renal failure, coagulopathy and endothelial dysfunction appear to be implicated (6). The SARS-CoV-2 virus gains entry to cells through binding to the ACE2 receptor which can be found in a variety of organs including the heart, in theory having the propensity to have a direct effect on the myocardium (7–9). Cardiac imaging studies of patients recovered from COVID-19 show evidence of a range of patterns and localisation of myocardial injury, with acute and sub-acute myocardial inflammation suggesting distinct pathways leading to myocardial damage (10).

Elevated cardiac biomarkers have been found to be associated with poorer outcomes in COVID-19. Studies have shown that the presence of elevated cardiac biomarkers including cardiac troponins appears to be associated with both death and increased intensive care admissions, as well as the requirement for ventilation (11). As well as biochemical markers, multiple demographic factors have been identified as adverse prognostic predictors in COVID-19 infection. These include increasing age, male sex, positive smoking status, comorbidities including diabetes and obesity, chronic major organ diseases and autoimmune diseases (12).

With the widespread administration and uptake of vaccinations in western nations the risk of severe COVID-19 has been significantly mitigated but not eliminated(13). This relates to the emergence of vaccine resistant variants, and endemicity of infection in the population in vulnerable and non-vaccinated individuals. COVID-19 infection will likely remain an ongoing public health issue with further surges and pandemics; continuing to characterise the clinical profile and prognostic features of patients with COVID-19 is important to identify those at higher risk of severe disease and hospitalisation. In this study, we primarily aimed to better delineate and clarify the relationship between elevated cardiac biomarkers and mortality in COVID-19 in a population of hospitalised COVID-19 patients at an inner-city London district general hospital during the first wave of the pandemic in early 2020. We also investigated other factors that may be implicated in the prognostication of COVID-19 patients.

## Methods

### Data collection

All adults admitted with swab-proven RT-PCR COVID-19 to Homerton University Hospital (HUH), London, from the date of the first positive swab from 02/03/2020 to 30/04/2020 were eligible for inclusion. Patients were identified using the hospital coding system applied to unselected adults admitted for acute medical care at HUH. Patients under the age of 18 were excluded and those who did not have a positive SARS-CoV-2 real time reverse-transcriptase polymerase chain reaction test (RT-qPCR) were excluded.

We retrospectively analysed the patient records from the electronic patient record (EPR) with the primary outcome defined as mortality within 28 days of admission to HUH. Data including observations and laboratory tests (including serum high sensitivity troponin I (hs-TnI)) were automatically extracted from the EPR. Additional data including admission ECG, presenting symptoms and outcomes were then manually collected for these patients from EPR and patient case notes where required. Manual validation of a random sample of 10% of the dataset was completed. The study was submitted to the HRA (Health Research Authority) and was approved by the HUH trust board.

### Patient and Public Involvement

Patients and the public were not involved in the design or analysis of the study.

### Laboratory methods

Serum hs-TnI levels were assessed by a high-sensitivity cardiac troponin I microparticle chemiluminescent immunoassay (ARCHITECT STAT, hs-TNI, Abbott) on the fully automated Abbott ARCHITECT analyser (Abbott ARCHITECT *STAT* high sensitive troponin-I. Package insert. G1-0139/R02. 2013). The upper reference limit (URL) of hs-TNI defined as the 99^th^ percentile of hs-TNI distribution in a reference population was 15.5 ng/l for females, and 34 ng/l for males respectively. Limit of blank and limit of detection have been determined as 0.7-1.3 ng/l and 1.1-1.9 ng/l respectively.

### Statistical analysis

Data were analysed according to the primary outcome of death during hospital admission. A Mann Whitney U Test was performed to compare troponin values against the primary outcome, and a Chi-squared test was performed to compare troponin positivity against the primary outcome. Further Chi-square, fisher’s exact and Mann Whitney U tests were performed to compare troponin positivity to demographic, laboratory, imaging, and other outcomes including ICU admission, but these were not corrected for multiple testing and should be interpreted as exploratory (tables 1-3). Confidence intervals were computed for the above testing. A p value < to 0.05 was considered as significant. Univariate and multivariate logistical regression analyses were performed to evaluate the link between troponin positivity and death. Numerous different demographics, clinical and biochemical variables were tested for association with short term mortality and area under the curve (AUC) was computed for all regressions. Sensitivity and specificity were computed for the multivariate regression. All data analysis was performed in the statistical computing software R (version 4.0.3). Statistical tests used were discussed with an independent statistician to ensure the appropriateness of the tests used.

**Table 1:** Demographics and patient characteristics. Note that where a multiclass comparator was made p-values (for example for gender) p-values are the same for all groups. Similarly, for some comparisons such as age, where the input data was continuous but has been converted to ordinal, the distributions of individual values within a group have been compared using a Mann-Whitney U test, and again the p-values are the same for all groups.

| Parameter | Troponin positive group (n, %) | Troponin negative group (n, %) | Chi-square P-value |
| --- | --- | --- | --- |
|  | Troponin positive (N=124) | Troponin negative (N=195) |  |
| *Gender* |  |  |  |
| Male | 58 (46.8) | 115 (59.0) | *P = 0.0437 |
| Female | 66 (53.2) | 80 (41.0) | *P = 0.0437 |
| *Age* |  |  |  |
| <18 | 0 (0) | 0 (0) | ***P<0.0001 (Mann-Whitney U) |
| 18-35 | 0 (0) | 20 (10.3) |  |
| 36-49 | 4 (3.2) | 25 (12.8) |  |
| 50-64 | 30 (24.2) | 83 (42.6) |  |
| ≥65 | 90 (72.6) | 67 (34.4) |  |
| *Ethnicity* |  |  |  |
| White | 38 (30.6) | 52 (26.7) | P = 0.709 |
| Asian | 7 (5.6) | 12 (6.2) |  |
| Black | 39 (31.5) | 64 (32.8) |  |
| Mixed | 3 (2.4) | 2 (1.0) |  |
| Other | 28 (22.6) | 55 (28.2) |  |
| Not declared | 9 (7.3) | 10 (5.1) |  |
| *Comorbidities* |  |  |  |
| Ischaemic heart disease | 28 (22.6) | 19 (9.7) | **P = 0.00278 |
| Heart Failure | 23 (18.5) | 8 (4.1) | ***P<0.0001 |
| Chronic Kidney Disease | 45 (36.3) | 13 (6.7) | ***P<0.0001 |
| Hypertension | 89 (71.8) | 93 (47.7) | ***P<0.0001 |
| Diabetes Mellitus | 56 (45.2) | 81 (41.5) | P = 0.602 |
| Dyslipidaemia | 45 (36.3) | 46 (23.6) | *P = 0.0203 |
| Lung disease | 31 (25) | 49 (25.1) | P = 1.00 |
| Other major organ disease | 80 (64.5) | 88 (45.1) | **P = 0.00109 |
| *Presenting complaint* |  |  |  |
| Chest pain | 7 (5.6) | 15 (7.7) | P = 0.634 |
| Shortness of Breath | 68 (54.8) | 113 (57.9) | P = 0.667 |
| Cough | 60 (48.4) | 104 (53.3) | P = 0.455 |
| Fever | 55 (44.4) | 99 (50.8) | P = 0.316 |
| *Smoking status* |  |  |  |
| Current smoker | 4 (3.2) | 14 (7.2) | *P = 0.00890 |
| Ex smoker | 26 (21) | 38 (19.5) | *P = 0.00890 |
| Never smoker | 76 (61.3) | 134 (68.7) | *P = 0.00890 |
| Not documented | 18 (14.5) | 9 (4.6) | *P = 0.00890 |
| *Baseline medications* |  |  |  |
| Statin therapy | 75 (60.4) | 79 (40.5) | **P = 0.000767 |
| ACEi/ARB | 85 (57.5) | 53 (27.2) | ***P<0.0001 |
| Antiplatelets | 41 (33.1) | 25 (12.8) | ***P<0.0001 |
| Anticoagulation | 16 (12.9) | 12 (6.2) | P = 0.06097 |
| Beta blockers | 45 (36.3) | 33 (16.9) | **P = 0.000151 |
| MRA | 8 (6.5) | 0 (0) | **P = 0.00126 |
| Other diuretic | 38 (30.6) | 25 (12.8) | **P = 0.0001741 |
| Other antihypertensive | 48 (38.7) | 64 (32.8) | P = 0.340 |

**Table 2:**
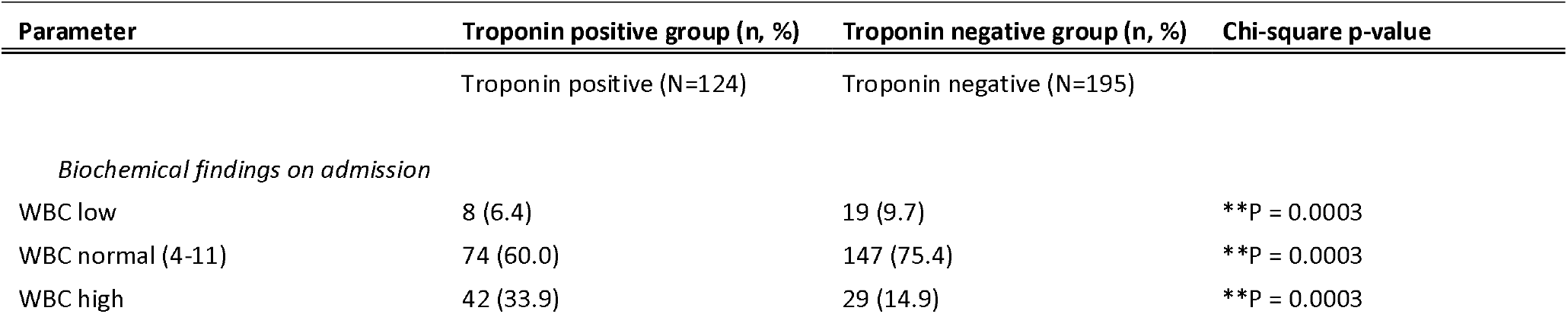
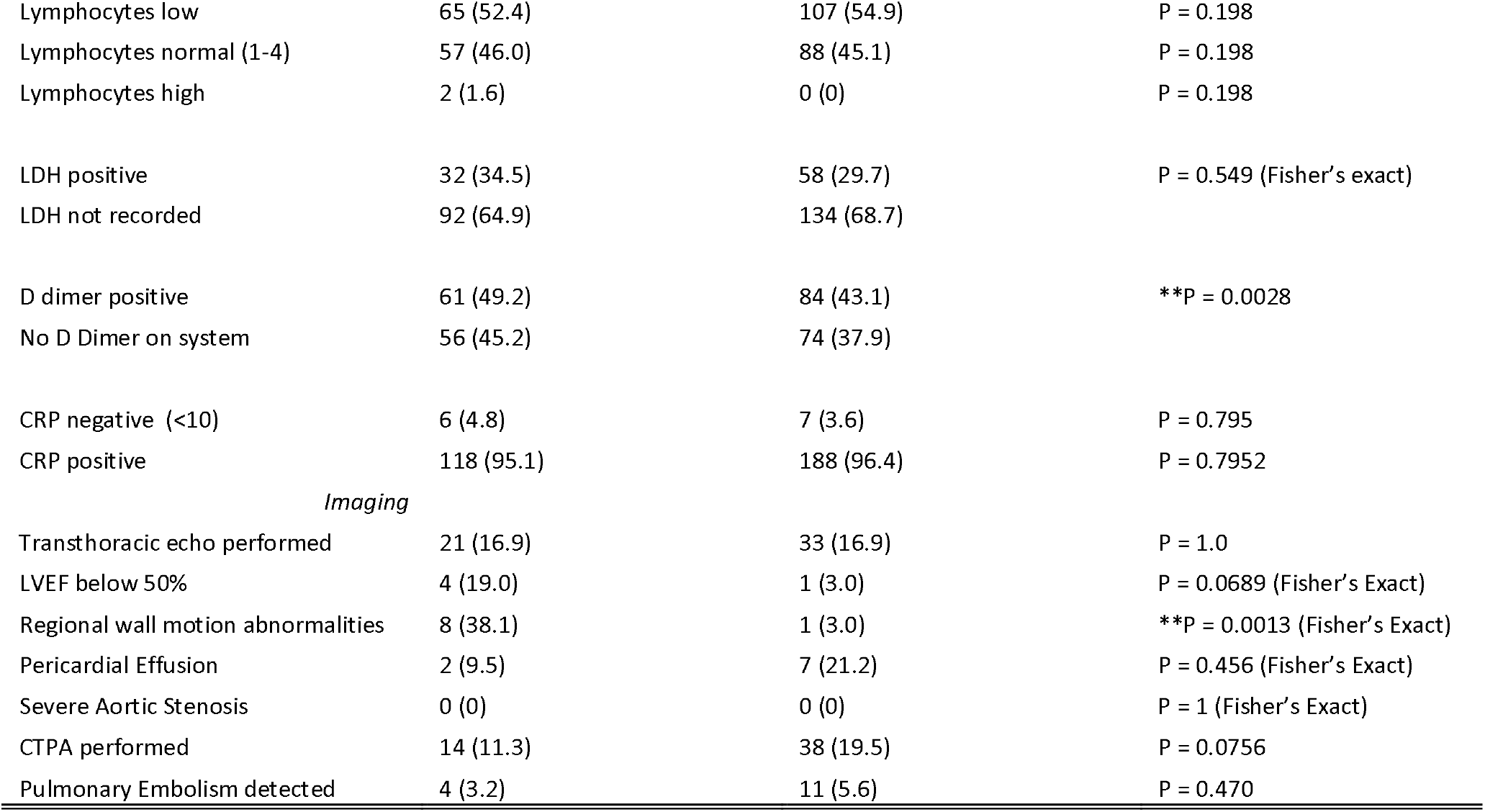
Inpatient investigations. Note that where a multiclass comparator was made p-values (for example for gender) p-values are the same for all groups. Similarly, for some comparisons such as age, where the input data was continuous but has been converted to ordinal, the distributions of individual values within a group have been compared using a Mann-Whitney U test, and again the p-values are the same for all groups.

**Table 3:** Outcomes and inpatient management

| Parameter | Troponin positive group, n,( %) | Troponin negative group, n(%) | Chi-square P-value |
| --- | --- | --- | --- |
|  | Troponin positive (N=124) | Troponin negative (N=195) |  |
| *Mortality* |  |  |  |
| Alive on discharge | 48 (38.7) | 145 (74.4) | ***P<0.0001 |
| Inpatient death | 66 (53.2) | 37 (19.0) | ***P<0.0001 |
| Death following readmission | 4 (3.2) | 0 (0) | ***P<0.0001 |
| *Critical care management* |  |  |  |
| CPAP | 21 (16.9) | 34 (17.4) | P = 1.0 |
| Intubation and ventilation | 15 (12.1) | 33 (16.9) | P = 0.310 |
| ITU admission | 17 (13.7) | 35 (17.9) | P = 0.399 |
| *Other in-hospital management* |  |  |  |
| Dual antiplatelet | 9 (7.3) | 2 (1.0) | *P = 0.0078 |
| Anticoagulation | 26 (21.0) | 42 (21.5) | P = 1.0 |
| Antiarrhythmic | 6 (4.8) | 8 (4.1) | P = 0.974 |
| Angioplasty/PCI | 1 (0.8) | 0 (0) | P = 0.819 |
| ACS Medical protocol | 7 (5.6) | 2 (1.0) | *P = 0.0373 |
| Immunosuppression | 8 (6.5) | 21 (10.8) | P = 0.268 |

## Results

### Population Characteristics

402 adult inpatients were found to have swab-proven RT-PCR COVID-19 between 02/03/2020 30/04/2020. 83 patients did not have a troponin measured and were therefore excluded. The baseline demographics and clinical characteristics for all patients are shown in table 1. There was a male preponderance to the troponin negative group with the inverse the case in the troponin positive group. Around one third of patients had a Black or Asian ethnicity. Hypertension was the most frequent recorded co-morbidity, followed by diabetes mellitus in our study population. Hypertension, hyperlipidaemia, heart failure, ischaemic heart disease and chronic kidney disease were significantly more prevalent in the troponin positive group. Higher use of medications in the troponin positive group such as ACE-I reflected the increased prevalence of these co-morbidities. Chest pain was uncommon in both groups. However, there was no significant difference in presenting symptom between the two groups.

### Inpatient investigations

High D-dimer, low lymphocyte count and high CRP was evident in both the troponin positive and troponin negative result (see table 2). 73% of patients over 65 years old had a positive troponin. There were a low number of computed tomography pulmonary angiograms (CTPAs) performed in this cohort. In addition, only around one third of troponin positive cases had a transthoracic echocardiogram completed. Reported abnormalities on imaging included reduced left ventricular ejection fraction, regional wall motion abnormalities and pericardial effusion.

### Outcomes and inpatient management

Table 3 shows the cumulative outcomes in the troponin positive and negative groups. The average age of patients who died was 74.0 (95 CI 71.5 – 76.5) years whilst the average age in those who survived was 60.4 (95 CI 58.3 – 62.5) years (see figure 1 panel C). Death rates were higher in the troponin positive group. Chi-squared test showed that survival of COVID-19 patients was significantly higher in those with a negative troponin compared to those with a positive troponin (p = 3.23 ×10^−10^). Mean initial troponin in patients who survived was 70.0ng/l (95 CI 5.6ng/l – 134.4ng/l), whilst in those who died it was 131.4ng/l (95 CI 78.9ng/l - 183.9ng/l) (see figure 1 panel D). A Mann Whitney U test showed that initial troponin was significantly higher in those who died (p = 2.24 ×10^−12^) compared to those who were alive. COVID-19 was the primary cause of death in both patient populations on the medical certificate for cause of death (MCCD). Only a minority of troponin positive patients had cardiovascular disease mentioned on the death certificate (2.4% in part 1, and 15.5% in part 2). A higher proportion of the troponin positive group received medical management for ACS, however only one patient subsequently went on to have percutaneous coronary intervention

**Figure 1.**
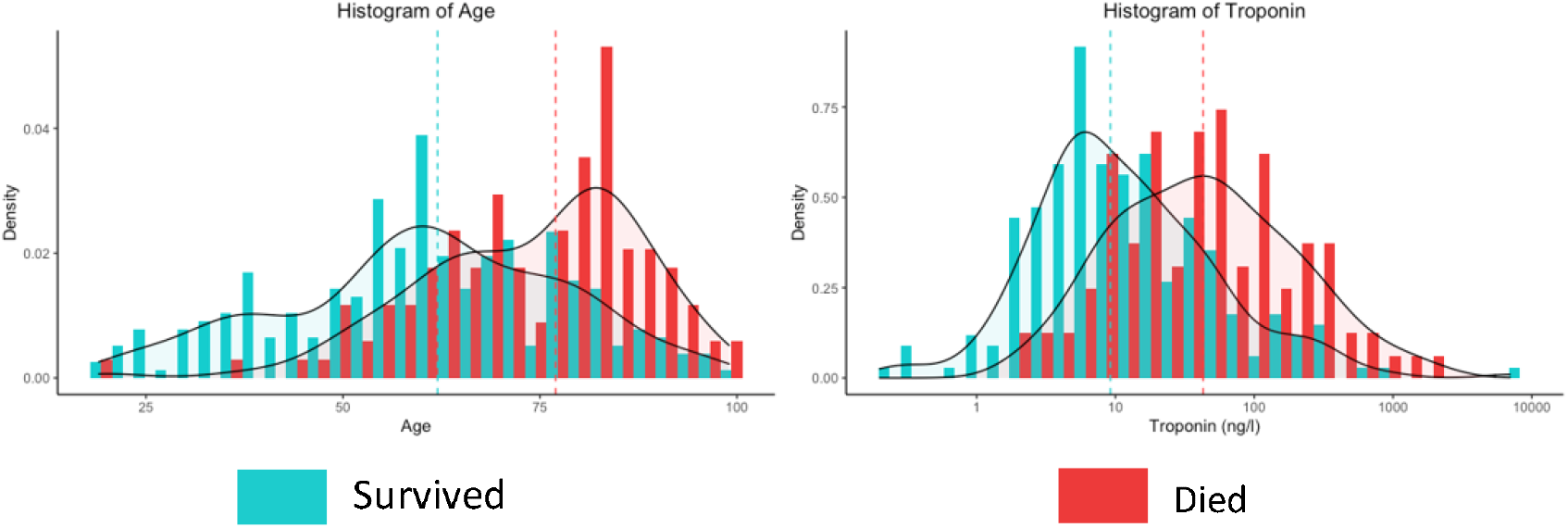
Histograms of age and troponin coloured by mortality. It is worth noting here that troponin is logarithmically scaled.

### Predictors of outcome

Table 4 shows the results of the univariate and multivariate regression in our patient cohort. A histogram of the odds ratio for the multivariate analysis can be seen in figure 2. In the multivariate logistical regression, lung disease, age, troponin positivity, and CPAP were all significantly associated with death, with an AUC of 0.893, sensitivity of 0.896 and specificity of 0.662 for the model. The receiver-operator (ROC) curve is shown in figure 3. Within this model, troponin positivity was independently associated with short term mortality (OR 2.96, 95% CI 1.34-6.71, p=0.0077).

**Table 4:**
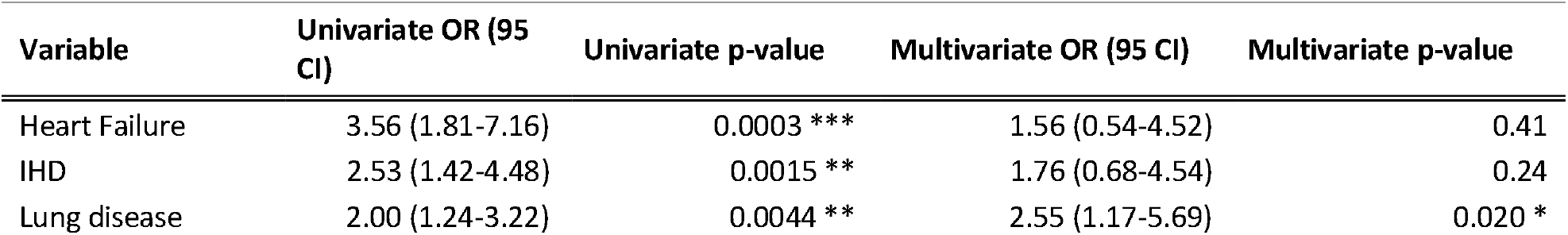
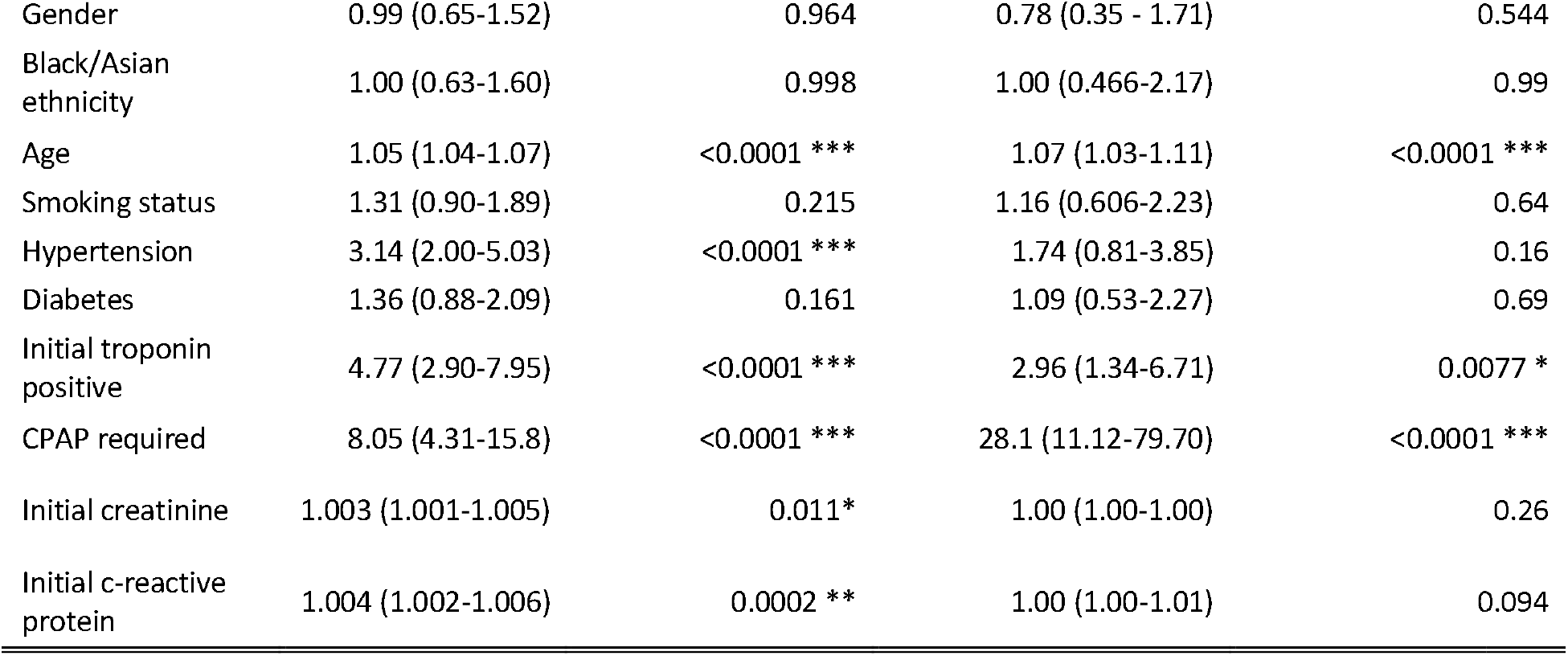
Multivariate and univariate analysis of predictors for mortality. Odds ratios and p-values generated using a logistic regression.

| Variable | Univariate OR (95 CI) | Univariate p-value | Multivariate OR (95 CI) | Multivariate p-value |
| --- | --- | --- | --- | --- |
| Heart Failure | 3.56 (1.81-7.16) | 0.0003 *** | 1.56 (0.54-4.52) | 0.41 |
| IHD | 2.53 (1.42-4.48) | 0.0015 ** | 1.76 (0.68-4.54) | 0.24 |
| Lung disease | 2.00 (1.24-3.22) | 0.0044 ** | 2.55 (1.17-5.69) | 0.020 * |
| Gender | 0.99 (0.65-1.52) | 0.964 | 0.78 (0.35 - 1.71) | 0.544 |
| Black/Asian ethnicity | 1.00 (0.63-1.60) | 0.998 | 1.00 (0.466-2.17) | 0.99 |
| Age | 1.05 (1.04-1.07) | <0.0001 *** | 1.07 (1.03-1.11) | <0.0001 *** |
| Smoking status | 1.31 (0.90-1.89) | 0.215 | 1.16 (0.606-2.23) | 0.64 |
| Hypertension | 3.14 (2.00-5.03) | <0.0001 *** | 1.74 (0.81-3.85) | 0.16 |
| Diabetes | 1.36 (0.88-2.09) | 0.161 | 1.09 (0.53-2.27) | 0.69 |
| Initial troponin positive | 4.77 (2.90-7.95) | <0.0001 *** | 2.96 (1.34-6.71) | 0.0077 * |
| CPAP required | 8.05 (4.31-15.8) | <0.0001 *** | 28.1 (11.12-79.70) | <0.0001 *** |
| Initial creatinine | 1.003 (1.001-1.005) | 0.011* | 1.00 (1.00-1.00) | 0.26 |
| Initial c-reactive protein | 1.004 (1.002-1.006) | 0.0002 ** | 1.00 (1.00-1.01) | 0.094 |

**Figure 2:**
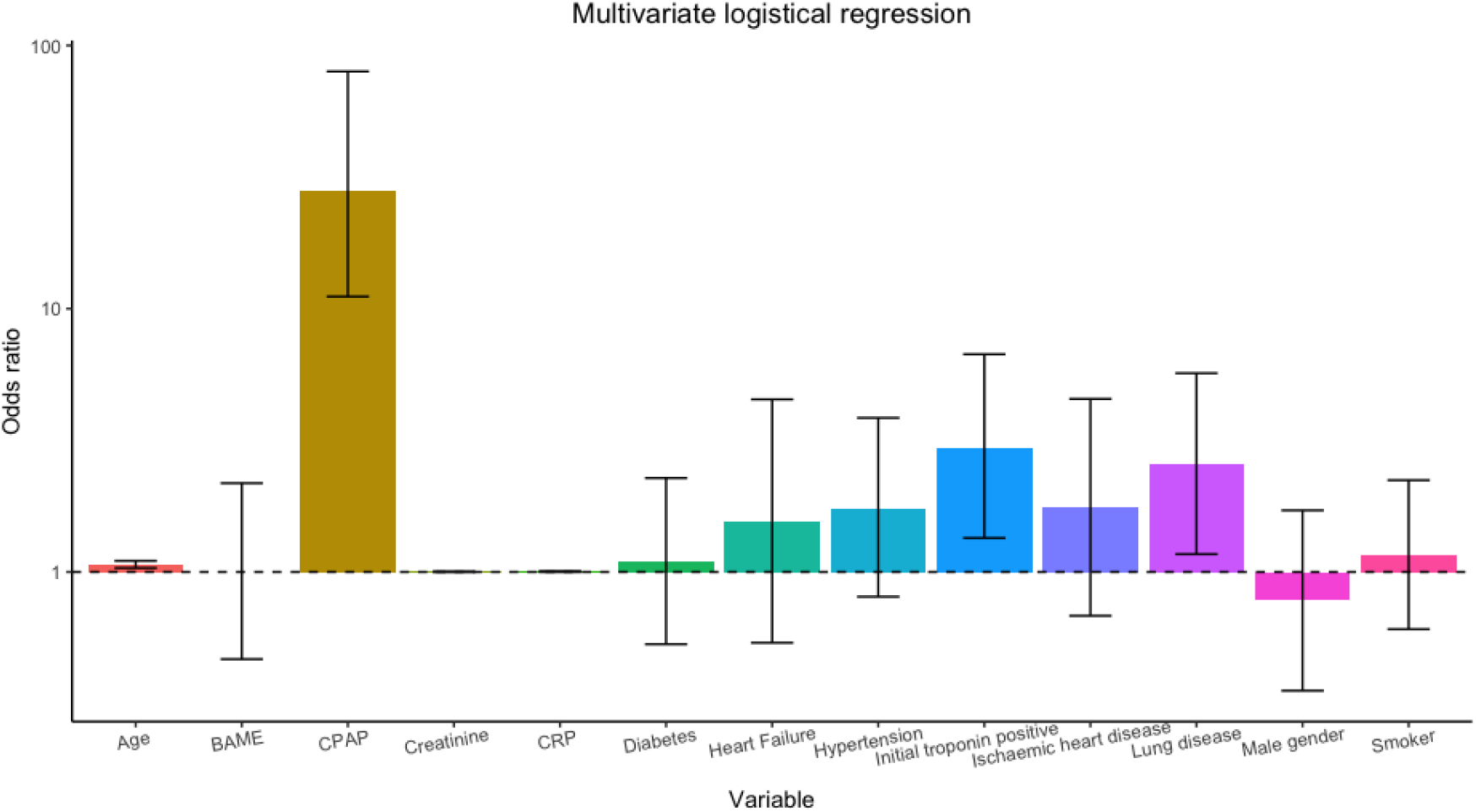
Odds ratios and associated p-values for input variables to logistic regression.

## Discussion

In this retrospective cross-sectional study, we have demonstrated that a positive initial troponin was independently associated with short-term mortality in patients hospitalized with COVID-19 These findings line up with other studies and meta-analyses (3–5,14–18,20,22,41–44). Papargeorgeiou et al. similarly reported the association between a positive initial troponin and mortality across several London hospitals covering different geographical areas (20). Other studies have found correlation between elevated troponin and more severe disease, ICU admission and the requirement for non-invasive and mechanical ventilation (3,4,14,22,42,43,45,46). We did not find any significant between group differences in CPAP use, ITU admission and intubation between troponin positive and negative groups. However, this observation based on relatively small numbers of patients and should be interpreted with caution. Troponin measurements appear to be helpful in identification of those at risk of death from COVID-19. Even in non-COVID hospitalised populations in the UK elevated troponin has been seen to be associated with mortality suggesting that myocardial injury forms part of the clinical picture in other non-cardiac severe illness (47).

In addition to troponin, increasing age, the need for CPAP and underlying lung disease were also independently associated with increased mortality in our cohort. Increasing mortality with increasing age is clearly correlated (48). Lung disease, in particular COPD and interstitial lung disease, has been shown to modestly increase the risk of severe COVID-19 in the UK, although asthma doesn’t appear to confer increased risk (49). The use of CPAP was associated with an increased risk of mortality in our cohort suggesting another interesting clinical marker to identify patients at higher risk of mortality.. The scope of this study was not to determine the effectiveness of CPAP; patients who require CPAP are clinically more unstable with severe disease and are more likely to deteriorate. The clear benefits of CPAP have been outlined in other studies designed as such (50).

In our cohort initial troponin was positive in 31% of patients. Data from Wuhan initially outlined an incidence of 12-27% (2–4). Subsequent studies from western countries show a broad prevalence of myocardial injury. In general, there appears to be higher frequency of troponin elevation in hospitalized patients in European and USA populations, when compared to China, with an incidence rising above 50% in some instances (11,14–21). A large meta-analysis including 49 studies from a combination of the USA, Europe and China showed an incidence of troponin elevation in 20.8% when measured in the first 24 hours, rising to 34.2% when measured during the ongoing hospital stay (5). Our finding appears to be broadly in line with the prevalence expected from the heterogenous evidence base.

Hypertension, chronic kidney disease, ischaemic heart disease and heart failure were all more common in the troponin positive group in our cohort, which is similarly seen in the large cohorts from the USA (18,22). These co-morbidities have been all been found to be associated with increased risk of death COVID-19 in large studies and similarly we found a higher prevalence in those that died (23,24). Dysfunction of the renin-angiotensin-aldosterone system (RAAS) may be implicated here. Binding of the virus to ACE2 with resultant loss of ACE2 may lead to increases in angiotensin II thereby contributing to endothelial dysfunction, vascular inflammation, and thrombosis (25). The dysfunctional RAAS seen in such co-morbidities may lead to susceptibility to severe disease via further disruption through viral binding of ACE2. A significant percentage of the patients in those with positive troponins had chronic kidney disease. A correlation in declining eGFR and rising serum troponin is known and may account for some of the elevated troponin levels. However, there was a broad spread of patients across chronic kidney disease groups 1-5, so it is unlikely to explain troponin elevation in the entirety of this group (26).

Severe COVID-19 is often characterised by a cytokine storm. The cytokine storm may be another factor implicated in myocardial injury as CRP levels have been seen to correlate with troponin levels (3). High levels of circulating cytokines may have a direct effect on the myocardium, as well as contributing to generalised endothelial dysfunction and coagulopathy (27). This prothrombotic state found in COVID-19 can contribute to various sequalae that can lead to myocardial strain and injury including pulmonary emboli (28). Pulmonary embolism has been reported in 13% of patients in a large systematic review, in our sample the prevalence of pulmonary embolism was rather lower, and prevalence was higher in the troponin negative group (29). However, the number of CT pulmonary angiography scans performed in our sample was also low so our results should be interpreted with caution.

Case series of occlusive coronary disease in COVID-19 exist in the literature, however it appears to be relatively uncommon (30,31). This is reflected in our sample with only 1 patient proceeding to invasive coronary angiogram and only 9 of the patients with positive troponin receiving medical ACS treatment during their admission. In ST elevation myocardial infarction associated with COVID-19 a higher thrombotic burden has been noted during angiography, suggesting that the prothrombotic state may be a contributing factor to coronary occlusion however clear causation is not yet proved (32,33). Case reports of clinical myocarditis do exist in the literature, however the prevalence in autopsy studies appears to be relatively low with one compilation of autopsy studies estimating histopathological evidence myocarditis at 7.2%, being only functionally significant in <2% (34–36). In our cohort we did not identify any cases of myocarditis. However, we acknowledge that myocarditis is recognised to rarely occur following COVID-19 infection and vaccination, but this association needs to be further investigated (37). Acute COVID-19 myocarditis has been detected on cardiac MRI but we were unable to correlate this in our study (10).

Only around a third of those with a positive troponin underwent echocardiography in our cohort. The prevalence of regional wall motion abnormalities, impaired left ventricular systolic function and pericardial effusion were proportionally higher in the troponin positive group. These findings have previously been characterised as part of the clinical picture of myocardial injury in COVID-19 (38–40). However, our results on echocardiographic findings should be approached with caution as the number of patients in the troponin positive group undergoing echocardiography was three times higher than that in the negative group.

There are some limitations to our findings. First, our sample was a small retrospective analysis of patients with COVID-19 requiring hospitalization. Second, within this cohort of patients, a significant proportion (approximately 20%)did not have blood troponin levelsmeasured and were excluded from the analysis. Furthermore, there were differences in timing of sampling of blood troponin between patients during their hospital stay.

## Conclusions

Troponin positivity was independently associated with increased short-term mortality in hospitalized patients with COVID-19 patients during the first wave of the pandemic. The mechanisms implicated in myocardial injury in COVID-19 are not fully understood and requires further investigation.

## Data Availability

Data are not available as they are clinically privileged. Access to scripts used for analysis of data are available on request.

## Bibliography

1. World Health Organization. WHO Coronavirus (COVID-19) Dashboard WHO Coronavirus (COVID-19) Dashboard With Vaccination Data [Internet]. 2021 [cited 2021 Sep 2]. Available from: https://covid19.who.int/

2. Huang C, Wang Y, Li X, Ren L, Zhao J, Hu Y, et al. Clinical features of patients infected with 2019 novel coronavirus in Wuhan, China. The Lancet. 2020;

3. Guo T, Fan Y, Chen M, Wu X, Zhang L, He T, et al. Cardiovascular Implications of Fatal Outcomes of Patients with Coronavirus Disease 2019 (COVID-19). JAMA Cardiology. 2020;

4. Shi S, Qin M, Shen B, Cai Y, Liu T, Yang F, et al. Association of Cardiac Injury with Mortality in Hospitalized Patients with COVID-19 in Wuhan, China. JAMA Cardiology. 2020;

5. Zhao BC, Liu WF, Lei SH, Zhou BW, Yang X, Huang TY, et al. Prevalence and prognostic value of elevated troponins in patients hospitalised for coronavirus disease 2019: a systematic review and meta-analysis. Journal of Intensive Care. 2020;

6. Imazio M, Klingel K, Kindermann I, Brucato A, De Rosa FG, Adler Y, et al. COVID-19 pandemic and troponin: Indirect myocardial injury, myocardial inflammation or myocarditis? Heart. 2020.

7. Hoffmann M, Kleine-Weber H, Schroeder S, Krüger N, Herrler T, Erichsen S, et al. SARS-CoV-2 Cell Entry Depends on ACE2 and TMPRSS2 and Is Blocked by a Clinically Proven Protease Inhibitor. Cell. 2020;

8. Zou X, Chen K, Zou J, Han P, Hao J, Han Z. Single-cell RNA-seq data analysis on the receptor ACE2 expression reveals the potential risk of different human organs vulnerable to 2019-nCoV infection. Frontiers of Medicine. 2020;

9. Chen L, Li X, Chen M, Feng Y, Xiong C. The ACE2 expression in human heart indicates new potential mechanism of heart injury among patients infected with SARS-CoV-2. Cardiovascular research. 2020;

10. Kotecha T, Knight DS, Razvi Y, Kumar K, Vimalesvaran K, Thornton G, et al. Patterns of myocardial injury in recovered troponin-positive COVID-19 patients assessed by cardiovascular magnetic resonance. European Heart Journal [Internet]. 2021 May 14 [cited 2021 Sep 1];42(19):1866–78. Available from: https://academic.oup.com/eurheartj/article/42/19/1866/6140994

11. Majure DT, Gruberg L, Saba SG, Kvasnovsky C, Hirsch JS, Jauhar R. Usefulness of Elevated Troponin to Predict Death in Patients With COVID-19 and Myocardial Injury. American Journal of Cardiology. 2020;

12. Izcovich A, Ragusa MA, Tortosa F, Marzio MAL, Agnoletti C, Bengolea A, et al. Prognostic factors for severity and mortality in patients infected with COVID-19: A systematic review. PLoS ONE [Internet]. 2020 Nov 1 [cited 2021 Sep 26];15(11). Available from: /pmc/articles/PMC7671522/

13. Public Health England. Direct and indirect impact of the vaccination programme on COVID-19 infections and mortality [Internet]. London; 2021 [cited 2021 Sep 2]. Available from: https://assets.publishing.service.gov.uk/government/uploads/system/uploads/attachment_data/file/997495/Impact_of_COVID-19_vaccine_on_infection_and_mortality.pdf

14. Shah P, Doshi R, Chenna A, Owens R, Cobb A, Ivey H, et al. Prognostic Value of Elevated Cardiac Troponin I in Hospitalized Covid-19 Patients. American Journal of Cardiology. 2020;

15. Poterucha TJ, Elias P, Jain SS, Sayer G, Redfors B, Burkhoff D, et al. Admission cardiac diagnostic testing with electrocardiography and troponin measurement prognosticates increased 30-day mortality in COVID-19. Journal of the American Heart Association. 2020;

16. Manocha KK, Kirzner J, Ying X, Yeo I, Peltzer B, Ang B, et al. Troponin and Other Biomarker Levels and Outcomes Among Patients Hospitalized with COVID-19: Derivation and Validation of the HA 2 T 2 COVID-19 Mortality Risk Score. Journal of the American Heart Association. 2020;

17. Cordeanu E, Duthil N, Severac F, Lambach H, Tousch J, Jambert L, et al. Prognostic Value of Troponin Elevation in COVID-19 Hospitalized Patients. Journal of clinical medicine. 2020;9(12).

18. Lala A, Johnson KW, Januzzi JL, Russak AJ, Paranjpe I, Richter F, et al. Prevalence and Impact of Myocardial Injury in Patients Hospitalized With COVID-19 Infection. Journal of the American College of Cardiology. 2020;

19. Arcari L, Luciani M, Cacciotti L, Musumeci MB, Spuntarelli V, Pistella E, et al. Incidence and determinants of high-sensitivity troponin and natriuretic peptides elevation at admission in hospitalized COVID-19 pneumonia patients. Internal and Emergency Medicine. 2020;

20. Lombardi CM, Carubelli V, Iorio A, Inciardi RM, Bellasi A, Canale C, et al. Association of Troponin Levels with Mortality in Italian Patients Hospitalized with Coronavirus Disease 2019: Results of a Multicenter Study. JAMA Cardiology. 2020;

21. Papageorgiou N, Sohrabi C, Merino DP, Tyrlis A, Atieh AE, Saberwal B, et al. High sensitivity troponin and COVID-19 outcomes. Acta Cardiologica [Internet]. 2021 [cited 2021 Sep 1];1. Available from: /pmc/articles/PMC7970632/

22. Majure DT, Gruberg L, Saba SG, Kvasnovsky C, Hirsch JS, Jauhar R. Usefulness of Elevated Troponin to Predict Death in Patients With COVID-19 and Myocardial Injury. American Journal of Cardiology. 2020;

23. Deng G, Yin M, Chen X, Zeng F. Clinical determinants for fatality of 44,672 patients with COVID-19. Critical Care. 2020.

24. Docherty AB, Harrison EM, Green CA, Hardwick HE, Pius R, Norman L, et al. Features of 20 133 UK patients in hospital with covid-19 using the ISARIC WHO Clinical Characterisation Protocol: Prospective observational cohort study. The BMJ. 2020;

25. South AM, Diz DI, Chappell MC. COVID-19, ACE2, and the cardiovascular consequences. American Journal of Physiology - Heart and Circulatory Physiology. 2020.

26. Chung JZY, Jones GRD. Effect of renal function on serum cardiac troponin T - Population and individual effects. Clinical Biochemistry. 2015;

27. Tersalvi G, Vicenzi M, Calabretta D, Biasco L, Pedrazzini G, Winterton D. Elevated Troponin in Patients With Coronavirus Disease 2019: Possible Mechanisms. Journal of Cardiac Failure. 2020.

28. Abou-Ismail MY, Diamond A, Kapoor S, Arafah Y, Nayak L. The hypercoagulable state in COVID-19: Incidence, pathophysiology, and management. Thrombosis Research. 2020.

29. Malas MB, Naazie IN, Elsayed N, Mathlouthi A, Marmor R, Clary B. Thromboembolism risk of COVID-19 is high and associated with a higher risk of mortality: A systematic review and meta-analysis. EClinicalMedicine. 2020;

30. Stefanini GG, Montorfano M, Trabattoni D, Andreini D, Andreini D, Ferrante G, et al. ST-Elevation Myocardial Infarction in Patients with COVID-19: Clinical and Angiographic Outcomes. Circulation. 2020;

31. Bangalore S, Sharma A, Slotwiner A, Yatskar L, Harari R, Shah B, et al. ST-Segment Elevation in Patients with Covid-19 — A Case Series. New England Journal of Medicine. 2020;

32. Little CD, Kotecha T, Candilio L, Jabbour RJ, Collins GB, Ahmed A, et al. COVID-19 pandemic and STEMI: Pathway activation and outcomes from the pan-London heart attack group. Open Heart. 2020;

33. Choudry FA, Hamshere SM, Rathod KS, Akhtar MM, Archbold RA, Guttmann OP, et al. High Thrombus Burden in Patients With COVID-19 Presenting With ST-Segment Elevation Myocardial Infarction. Journal of the American College of Cardiology. 2020;

34. Halushka MK, Vander Heide RS. Myocarditis is rare in COVID-19 autopsies: cardiovascular findings across 277 postmortem examinations. Cardiovascular Pathology. 2021;

35. Inciardi RM, Lupi L, Zaccone G, Italia L, Raffo M, Tomasoni D, et al. Cardiac Involvement in a Patient with Coronavirus Disease 2019 (COVID-19). JAMA Cardiology. 2020;

36. Wei X, Fang Y, Hu H. Glucocorticoid and immunoglobulin to treat viral fulminant myocarditis. European Heart Journal. 2020.

37. Kim HW, Jenista ER, Wendell DC, Azevedo CF, Campbell MJ, Darty SN, et al. Patients With Acute Myocarditis Following mRNA COVID-19 Vaccination. JAMA Cardiology [Internet]. 2021 [cited 2021 Sep 2]; Available from: https://jamanetwork.com/journals/jamacardiology/fullarticle/2781602

38. Giustino G, Croft LB, Stefanini GG, Bragato R, Silbiger JJ, Vicenzi M, et al. Characterization of Myocardial Injury in Patients With COVID-19. Journal of the American College of Cardiology. 2020;

39. Sud K, Vogel B, Bohra C, Garg V, Talebi S, Lerakis S, et al. Echocardiographic Findings in Patients with COVID-19 with Significant Myocardial Injury. Journal of the American Society of Echocardiography. 2020.

40. Rodríguez-Santamarta M, Minguito-Carazo C, Echarte-Morales J, Del Castillo-García S, Valdivia-Ruiz J, Fernández-Vázquez F. Echocardiographic findings in critical patients with COVID-19. Rev Española Cardiol. 2020;73(10):861–3.

41. Parohan M, Yaghoubi S, Seraji A. Cardiac injury is associated with severe outcome and death in patients with Coronavirus disease 2019 (COVID-19) infection: A systematic review and meta-analysis of observational studies. European Heart Journal: Acute Cardiovascular Care. 2020;

42. Li JW, Han TW, Woodward M, Anderson CS, Zhou H, Chen YD, et al. The impact of 2019 novel coronavirus on heart injury: A Systematic review and Meta-analysis. Progress in Cardiovascular Diseases. 2020.

43. Singh N, Anchan RK, Besser SA, Belkin MN, Cruz MD, Lee L, et al. High sensitivity Troponin-T for prediction of adverse events in patients with COVID-19. Biomarkers. 2020;

44. Salvatici M, Barbieri B, Cioffi S, Morenghi E, Leone F, Maura F, et al. Association between cardiac troponin I and mortality in patients with COVID-19. Biomarkers. 2020;25(8):634–40.

45. Lippi G, Lavie CJ, Sanchis-Gomar F. Cardiac troponin I in patients with coronavirus disease 2019 (COVID-19): Evidence from a meta-analysis. Progress in Cardiovascular Diseases. 2020.

46. Goudot G, Chocron R, Augy JL, Gendron N, Khider L, Debuc B, et al. Predictive Factor for COVID-19 Worsening: Insights for High-Sensitivity Troponin and D-Dimer and Correlation With Right Ventricular Afterload. Frontiers in Medicine. 2020;

47. Kaura A, Panoulas V, Glampson B, Davies J, Mulla A, Woods K, et al. Association of troponin level and age with mortality in 250 000 patients: Cohort study across five UK acute care centres. The BMJ. 2019;

48. Levin AT, Hanage WP, Owusu-Boaitey N, Cochran KB, Walsh SP, Meyerowitz-Katz G. Assessing the age specificity of infection fatality rates for COVID-19: systematic review, meta-analysis, and public policy implications. European Journal of Epidemiology 2020 35:12 [Internet]. 2020 Dec 8 [cited 2021 Sep 2];35(12):1123–38. Available from: https://link.springer.com/article/10.1007/s10654-020-00698-1

49. Aveyard P, Gao M, Lindson N, Hartmann-Boyce J, Watkinson P, Young D, et al. Association between pre-existing respiratory disease and its treatment, and severe COVID-19: a population cohort study. The Lancet Respiratory Medicine [Internet]. 2021 Aug 1 [cited 2021 Sep 2];9(8):909–23. Available from: http://www.thelancet.com/article/S2213260021000953/fulltext

50. Perkins GD, Ji C, Connolly BA, Couper K, Lall R, Baillie JK, et al. An adaptive randomized controlled trial of non-invasive respiratory strategies in acute respiratory failure patients with COVID-19. medRxiv [Internet]. 2021 Aug 4 [cited 2021 Sep 2];2021.08.02.21261379. Available from: https://www.medrxiv.org/content/10.1101/2021.08.02.21261379v1

51. Guragai N, Vasudev R, Hosein K, Habib H, Patel B, Kaur P, et al. Does Baseline Diuretics Use Affect Prognosis in Patients With COVID-19? Cureus [Internet]. 2021 Jun 10 [cited 2021 Sep 2];13(6). Available from: /pmc/articles/PMC8272599/

52. Pengo M, Stefanini G, Pivato C, Soranna D, Zambra G, Zambon A, et al. IN-HOSPITAL DIURETIC USE IS ASSOCIATED WITH WORSE OUTCOME IN PATIENTS WITH COVID-19. Journal of Hypertension [Internet]. 2021 Apr [cited 2021 Sep 2];39(Supplement 1):e38. Available from: https://journals.lww.com/jhypertension/Fulltext/2021/04001/IN_HOSPITAL_DIURETIC_USE_IS_ASSOCIATED_WITH_WORSE.101.aspx

